# Multi-ancestry epigenome wide association study of generalized anxiety disorder

**DOI:** 10.1101/2025.11.04.25339500

**Authors:** Priya Gupta, Shengxin Liu, Marco Galimberti, Sarah Beck, Keyrun Adhikari, Cecilia Dao, Yaira Z Nunez, Cassie Overstreet, Uri Bright, VA Million Veteran Program, Murray B. Stein, Gita A Pathak, Xueyi Shen, Joel Gelernter, Daniel F. Levey

## Abstract

Epigenetic studies face persistent challenges related to small sample sizes, particularly when using epigenome-wide array technologies. Presumably it is this limitation that has hindered the discovery of replicable and robust findings, much like the early struggles of genome-wide association studies. To address this gap, we conducted one of the largest epigenetic investigations of generalized anxiety disorder (GAD) using 43,504 participant’s data from the Million Veteran Program. Our analysis assessed differential DNA methylation between GAD cases and controls across three major genetic ancestry groups: European (EUR), African (AFR), and Latin American (AMR). We identified 49 CpG sites reaching epigenome-wide significance across these ancestries. However, when controlling for smoking either by adjusting for smoking status or restricting analyses to non-smokers in the EUR group only 2 and 5 significant CpG sites remained significant, respectively. To explore the predictive utility of these findings, we constructed methylation risk scores using a clumping and correlation method. The scores showed significant association with GAD phenotype in the leave-one out cross-validation within the MVP cohort but failed to replicate the association in an independent sample from Scotland. This may reflect insufficient power in the follow-up cohort, the effects of unmeasured confounding variables, or other unmeasured heterogeneity. Our findings underscore both the promise and the ongoing limitations of large-scale epigenetic studies, particularly the need for replication efforts and improved control of environmental confounders. Continued expansion of sample sizes, stratified analyses by ancestry, and careful consideration of lifestyle-related and other covariates will be essential in advancing the reliability and interpretability of peripheral epigenetic markers in psychiatric phenotypes like GAD.

## Introduction

While anxiety is a normal and often adaptive part of everyday life, when it becomes persistent and excessive it may become maladaptive. Generalized anxiety disorder (GAD) was first formalized as a distinct diagnosis in 1980 with the publication of the third edition of the Diagnostic and Statistical Manual of Mental Disorders III [1]. Here it was described as “a disorder of uncontrollable and diffuse anxiety or worry that is excessive or unrealistic” for at least 1 month. The definition has undergone relatively modest but meaningful refinements since then, most notably extending duration of symptoms (6 months or more), functional impairment, and improving differentiation from other anxiety and mood disorders.

Epigenetic mechanisms have been hypothesized to be important in the context of GAD, particularly changes in DNA methylation, histone modifications, and non-coding RNAs. DNA methylation of stress-related genes, such as retinoic acid receptor α *(RARA)* is a biologically plausible mechanism in the context of stress-related psychopathology.[3] While research on histone modifications in GAD is limited, these modifications are hypothesized to regulate the expression of stress-responsive genes. Although epigenome-wide association studies have not been previously conducted for GAD, there have been several studies in commonly co-morbid traits such as major depressive disorder[4] and post-traumatic stress disorder[5–7].

The clinical and genetic heterogeneity of GAD complicates efforts to pinpoint specific genetic or epigenetic contributors. Additionally, GAD-specific studies are underrepresented compared to other psychiatric disorders,[8] resulting in gaps in understanding regarding the unique genetic and epigenetic underpinnings.

Adequate sample size has been a challenge for epigenetic studies, including in use of epigenome-wide array technologies. Similar to the early days of Genome-Wide Association Studies (GWAS), where robust and replicable findings were difficult to detect, small sample sizes have been identified as one potential challenge to identifying robust epigenetic changes.[9] We present here a large epigenetic study of differential methylation in GAD, using data from the Million Veteran Program (MVP) using microarray analysis of peripheral blood-extracted DNA. We looked for differential methylation status between GAD cases and controls, stratified into three genetic ancestral groups: European (EUR), African (AFR) and Latin American (AMR). We tested the potential of methylation risk scores using clumping and correlation approach at capturing the GAD phenotype and saw significant association of methylation risk scores (MRS) in holdout samples within MVP. A relatively small independent samples of cases from Generation Scotland did not show similar evidence of replication, which could be indicative of power levels not yet sufficient to break free of “winner’s curse,” possible complications due to confounding factors in the independent sample replication, or other sources of heterogeneity.

## METHODS

### DNA Methylation Quantification

The Illumina Infinium Human Methylation Epic chip array v1 with 865,918 probes was run on the 45,500 available whole blood samples from MVP cohort to extract the DNA methylation level information. The signal intensities were converted into beta values from the IDAT files and then background correction was performed using the SeSAMe package [10]. During the probes-level QC, probes with high probability of out of band hybridization (pOOBAH) of more than 0.05 in more than 10% of samples were excluded. In sample-level QC, samples with less than 96% probes passing the pOOBAH p-value threshold of less than 0.05 were removed. Further, technically unreliable probes including probes with poor mapping quality, poor titration correlation, color channel switching, cross-hybridization were removed leaving 768,569 probes in the final dataset.

### GAD cases definition

We queried electronic health record (EHR) information of MVP participants to identify inpatient and outpatient visits using the VA Informatics and Computing Infrastructure (VINCI). We searched for ICD codes F41.1 and 300.02 (Generalized Anxiety Disorder diagnosis codes from ICD10 and ICD9, respectively), using methods reported previously [11]. Participants with at least one lifetime occurrence of these codes were classified as cases, those without were classified as controls.

### EWAS

Genetic ancestry of the samples was mapped using the genetic principal components generated from SNP-array based genotype data in MVP and comparing them to the 1000 Genomes reference panel as described in Hang *et al* studies [12, 13]. Meffil package was used to run EWAS [14]. Beta values were regressed against the binary GAD case/control status. Ancestry principal components (PC) were calculated from DNA methylation data. To leverage the variability arising due to technical bias, technical principal components were calculated using background and control signals probes. Blood cell composition in the samples (CD8+T, CD4+T, natural killer, B cells, monocytes, and neutrophils) was estimated using Houseman’s constrained projection algorithm [15]. Age, sex, Houseman cell type fraction, scanner ID used for assay, first 20 genomic PC (to leverage population stratification), first 20 technical PC were used as covariates to perform the regression. Quantile-quantile plots are shown in Suppl. Fig 1 & Supplementary Fig S1. BACON adjustment was applied to correct for bias and inflation [16].

### Sensitivity analysis

Life’s simple seven (LSS) survey responses were used to obtain the smoking and EHR was used to pull BMI information of the participants. Smoking status of each participant was binned into three possible states: current, former (did not smoke in last 12 months), or never (did not smoke for more than last 12 months). Categorical numerical values of 0,1 and 2 were used as scores to define the categories of smoking. These BMI and smoking values were then used as covariates in addition to previously described covariates for sensitivity analysis. The sensitivity EWAS was run for 26,099 samples of the EUR EWAS sample due to lack of BMI and smoking information for the remaining samples. A separate EWAS with only ‘never’ smoker samples was also performed (Figure S2).

### Blood-brain DNAm correlations

The BECon database was used to extract the blood-brain correlations of the CpG sites [17]. For each site, it provides the correlations values between blood and three brain tissues-Brodmann areas BA7, BA10 and BA20. Out of the 3 brain tissues, the tissue with highest absolute correlation value with blood was used to infer the brain-blood correlations. A rho value of >0.1 and <-0.1 was used to report the positively and negatively correlated sites respectively. The BECon provided blood-brain correlation values for 14 of the 33 identified sites. The remaining 19 sites are missing in the BECon database as they were not available on the 450k array or otherwise not reported.

### Gene set enrichment analysis

Gene set enrichment analysis (GSEA) was carried out using Gene2Func utility of FUMA. CpG sites were mapped to their respective genes using the Human Infinium MethylationEPIC BeadChip v1 manifest file. Genes with CpG sites showing epigenome-wide significant (EWS) GAD association were provided as input to the FUMA software and ‘All’ including non-coding regions were selected as background gene set. Bonferroni-corrected P-value of less than 0.05 was used to define the significant enrichment. Because statistical power was limited to detect all potentially relevant sites, we applied a relaxed P-value threshold (5e-05) to define a secondary set of associated CpG sites and corresponding gene sets for each EWAS, which were then evaluated in sensitivity analyses.

### Methylation risk scores

To calculate MRS, the discovery and validation datasets were defined from the EUR cohort: we randomly assigned 75% of the EUR samples as the discovery set and the remaining samples formed the validation set. GAD EWAS was carried out in the discovery set and beta values from the resulting summary statistics were used as weights to compute the MRS for the samples in the validation set. The MRS of a sample is calculated as the weighed sum of beta values of selected CpG sites. Different exponentially decreasing P-value cutoffs ranging from 0.005 to 5e-11, decreasing by an order of magnitude at each step, were used to select CpG sites from the discovery summary statistics and co-methylation signal were used as an extra filtering step for CpG sites selection in computing MRS. A co-methylation matrix was generated from the discovery data set and only one of CpG sites of any pair of CpG sites sharing a correlation coefficient of value greater than 0.3 was used.

We attempted to replicate our findings in Generation Scotland,[18] a large family-based health and wellbeing study aiming to understand the genetic, lifestyle, and environmental factors influencing physical and mental health. 16,516 of these individuals have methylation data from the Illumina Infinium Human Methylation Epi chip array (850k). We investigated GAD diagnosis within this cohort using ICD codes, similar to those used in the MVP. We identified 153 cases and 16,363 controls. We tested MRS using similar parameters as described above, except that testing P-value thresholds from MVP summary data of 0.05, 0.01, 0.005, and 0.001 were used. Receiver operating characteristics were calculated, from which the area under the curve (AUC) was generated to assess performance.

## RESULTS

### Ancestry-stratified GAD EWAS

We performed GAD EWAS in 3-different ancestries - European (EUR), African (AFR) and Admixed (AMR) in the MVP cohort using whole blood methylation data. Global genetic ancestry was assigned using 1000 Genomes Project Phase 3 EUR, AFR, and AMR reference panel and ten principal components (PCs) from within each stratified ancestry using genotype data, as previously described [11, 12, 19]. EUR EWAS was performed in n=27,783 samples including 1712 cases (Table 1). A lambda inflation factor value of 1.03 was observed for the EWAS. 33 EWS CpG sites (P<5.88e-08) on 13 different chromosomes were found to be associated GAD (**see Fig1A**). The top significant CpG sites mapped to genes including *AHRR, RARA, FURIN, ITPK1, CSRNP1, SUGT1P1*. Two of these EWS sites are part of a CpG island located on chr2 (chr2:233,283,397-233,285,959) and 11 sites are located on shores of 8 CpG islands on 8 chromosomes (Chr1:1,709,394-1,710,582, Chr3 :39,194,241-39,195,359, Chr5:373,842-374,426, Chr9:33,447,446-33,447824, Chr11:86,511,184-86,511,889, Chr15 :91,414,311-91,415,427 , Chr 17: 38,474,197-38,474,980 & Chr19: 17,000,627-17,001,398) suggesting these islands may have important regulatory roles in GAD gene biology. All the sites except the cg16552271 in *KIAA1026* showed negative beta, indicating that most sites were hypomethylated in GAD cases.

**Table 1.**
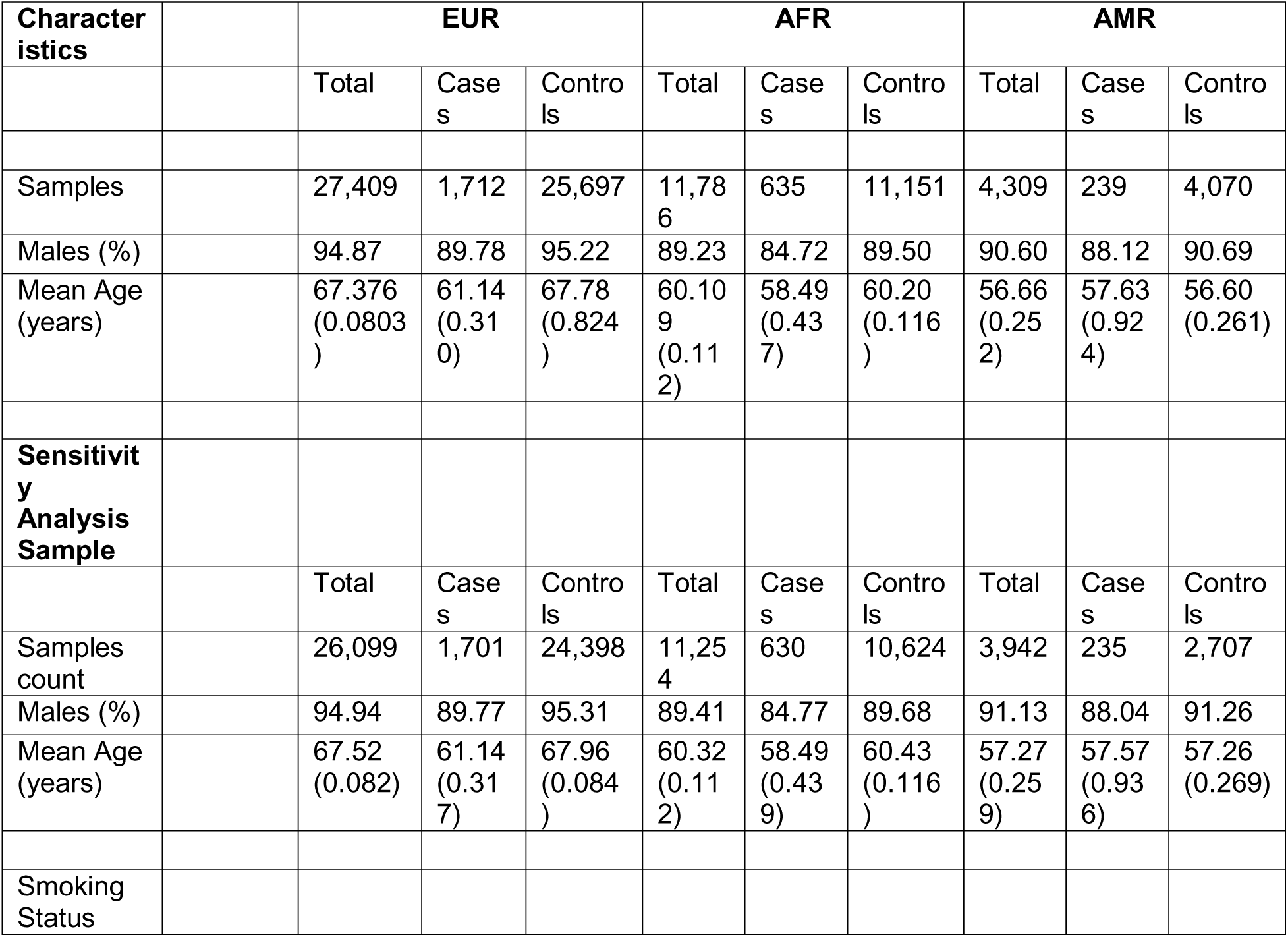
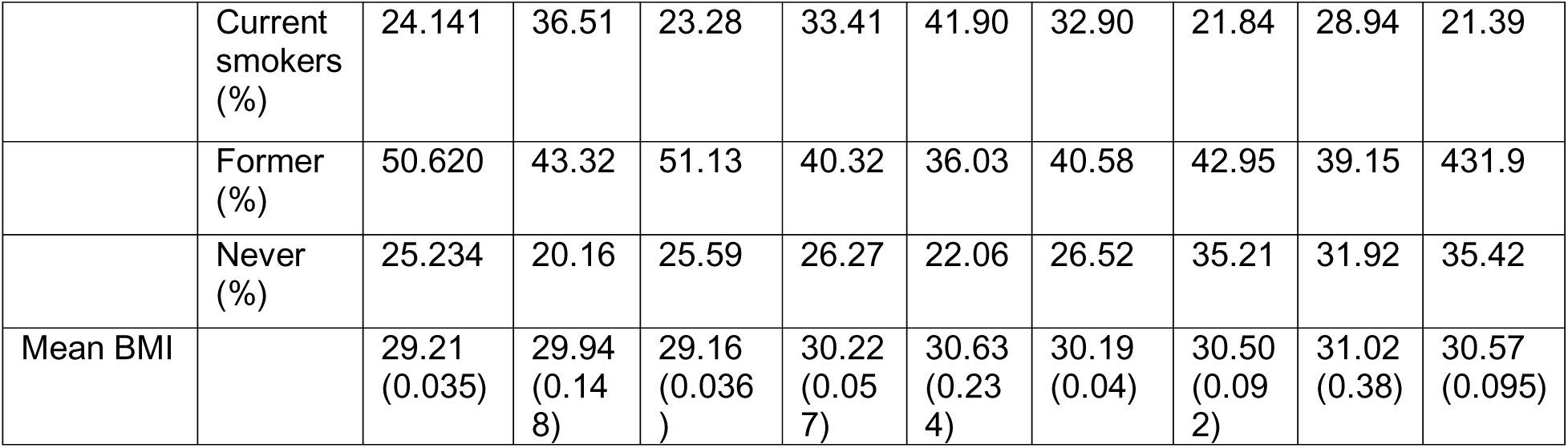
Population characteristics.

The AFR EWAS identified 9 EWS CpG sites where methylation status were associated with GAD (see **Fig2A**). Intergenic site Cg16962558 on chromosome 6 is the top-scoring site (P=2.24e-09). The remaining 8 sites mapped to 9 genes including *KRAS, ARHGAP12, HSDL2* (Table 2).

**Table 2:** GAD EUR Genes listed when the site is located within a gene.

| Probe ID | CHR | Position | Coefficient | Coefficient SE | P-value | Gene |
| --- | --- | --- | --- | --- | --- | --- |
| cg16552271 | 1 | 15392907 | 0.006510763 | 0.001251477 | 2.4508E-08 | KIAA1026 |
| cg09182189 | 1 | 1709203 | -0.003714198 | 0.000675821 | 6.1085E-10 | NADK |
| cg01895164 | 1 | 26332153 | -0.00516551 | 0.001056239 | 3.3530E-08 |  |
| cg04885881 | 1 | 11123118 | -0.006979358 | 0.001380436 | 1.1713E-08 |  |
| cg21566642 | 2 | 2.33E+08 | -0.012662717 | 0.002162221 | 4.5781E-11 |  |
| cg01940273 | 2 | 2.33E+08 | -0.007364614 | 0.001420448 | 5.0509E-09 |  |
| cg00501876 | 3 | 39193251 | -0.004129824 | 0.000845735 | 3.5108E-08 | CSRNP1 |
| cg16414530 | 3 | 66437745 | 0.004371895 | 0.000845436 | 2.9849E-08 | LRIG1 |
| cg05575921 | 5 | 373378 | -0.023027555 | 0.003246135 | 1.9442E-15 | AHRR |
| cg25648203 | 5 | 395444 | -0.007238828 | 0.001088804 | 8.9834E-14 | AHRR |
| cg23576855 | 5 | 373299 | -0.022593077 | 0.003819747 | 2.9642E-11 | AHRR |
| cg24859433 | 6 | 30720203 | -0.005866079 | 0.000992274 | 3.0327E-11 |  |
| cg15342087 | 6 | 30720209 | -0.004457196 | 0.000868609 | 7.1706E-09 |  |
| cg12009405 | 6 | 1.49E+08 | -0.00952952 | 0.001973579 | 4.9296E-08 |  |
| cg01010073 | 7 | 45065573 | -0.006686702 | 0.001256354 | 2.0125E-09 | CCM2 |
| cg15246238 | 7 | 5635134 | -0.006344956 | 0.00130695 | 4.1893E-08 | FSCN1 |
| cg09511513 | 8 | 20060014 | -0.00840767 | 0.001654287 | 9.8711E-09 | ATP6V1B2 |
| cg15167811 | 9 | 1.15E+08 | -0.008353941 | 0.001728639 | 4.8054E-08 | PTBP3 |
| cg02716826 | 9 | 33447032 | -0.005350323 | 0.001003147 | 1.8651E-09 | SUGT1P1;AQP3 |
| cg01435315 | 10 | 72348420 | -0.009001578 | 0.001749394 | 6.5367E-09 |  |
| cg25161899 | 11 | 85028237 | -0.008476897 | 0.001715202 | 2.4215E-08 | DLG2 |
| cg14262884 | 11 | 43757533 | -0.00623292 | 0.001230926 | 1.1145E-08 | HSD17B12 |
| cg03461577 | 11 | 33901166 | -0.005930785 | 0.001196202 | 2.1910E-08 | LMO2 |
| cg11660018 | 11 | 86510915 | -0.006047473 | 0.001208482 | 1.6329E-08 | PRSS23 |
| cg14391737 | 11 | 86513429 | -0.0096459 | 0.001913221 | 1.2836E-08 | PRSS23 |
| cg05284742 | 14 | 93552128 | -0.004778187 | 0.000894207 | 1.7433E-09 | ITPK1 |
| cg07162861 | 15 | 91416008 | -0.014048173 | 0.002743618 | 7.7106E-09 | FURIN |
| cg12722937 | 15 | 91416047 | -0.003414637 | 0.000674939 | 1.1468E-08 | FURIN |
| cg19572487 | 17 | 38476024 | -0.007048582 | 0.001467154 | 5.7271E-08 | RARA |
| cg17739917 | 17 | 38477572 | -0.012222315 | 0.00160465 | 1.5964E-17 | RARA |
| cg14129477 | 17 | 53422220 | -0.007729514 | 0.001584057 | 3.5896E-08 |  |
| cg21911711 | 19 | 16998668 | -0.008034605 | 0.001414801 | 1.6699E-10 | F2RL3 |
| cg03636183 | 19 | 17000585 | -0.010284156 | 0.001634752 | 1.6416E-12 | F2RL3 |

Five EWS sites were identified in AMR EWAS mapping to genes-*ZBTB2, LY75,* and *PLXNC1* (see **Fig2B**). In both the AFR and AMR analyses the plurality of associated CpG sites indicated *hyper*methylation in GAD cases, in contrast to our findings in EUR cases.

### Sensitivity analysis

DNA methylation is impacted by numerous factors including smoking and BMI [20, 21]. Therefore, to assess potential confounding of smoking or BMI associated CpG sites in our GAD EWAS, we performed a sensitivity EWAS where available smoking and BMI information was used as additional covariates to regress out the effects of these in GAD-methylation regression. Based on the availability of smoking/BMI information of participants, the sensitivity EWAS was performed in a slightly smaller samples than primary EWAS for all 3 ancestries (Table 1). In the smoking and BMI controlled GAD EWAS of EUR, two CpG sites showed epigenome-wide significance (Figure 3 and Table S1). One site maps to *LRIG1* on chr3 and second maps to *RARA* on chr17. Both of these sites were also EWS in the primary EUR analysis. In AFR, 7 out of the 9 EWS CpG sites in the primary EWAS remained EWS in the sensitivity EWAS as well, while the remaining two CpG sites showed only slightly higher P-values (cg25271375 with a P-value = 6.62e-08, cg07038191 with a P-value =7.24e-08) (see Fig3).

**Figure 1:**
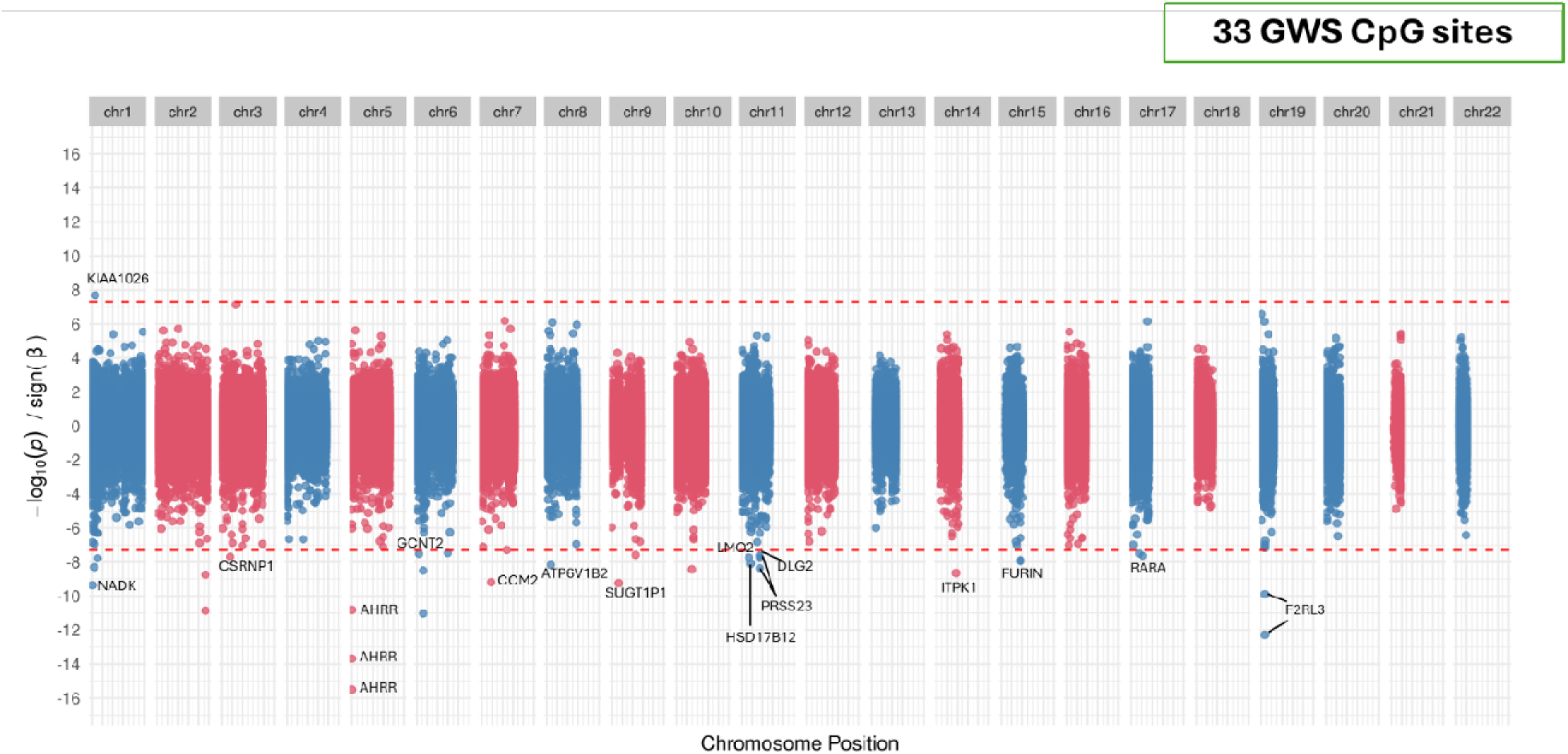
GAD EWAS in European ancestry. Miami plot of EUR EWAS.

**Figure 2:**
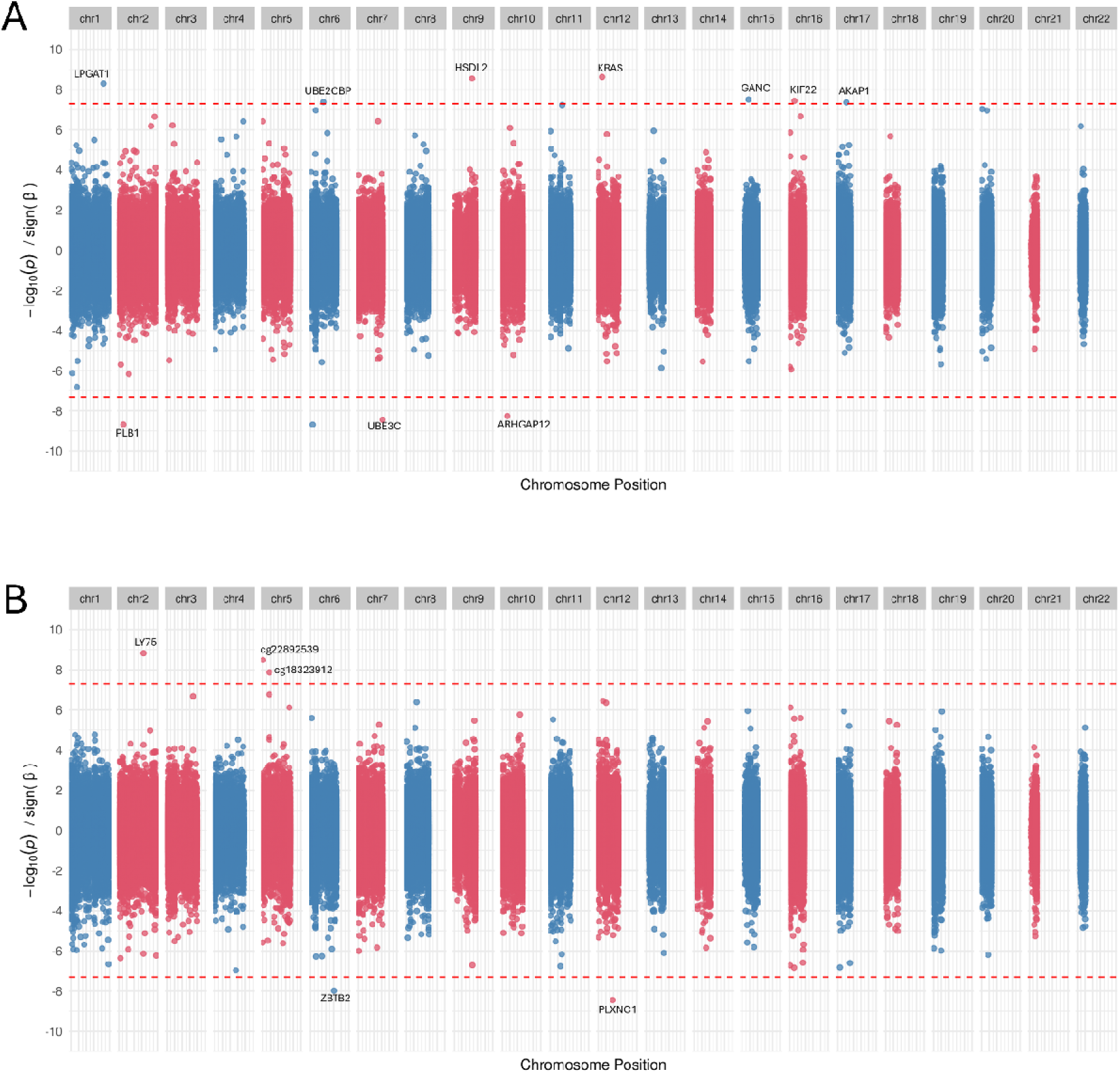
GAD EWAS in non-European ancestry. The figures displays the Miami plot of GAD EWAS in AFR (A) and AMR ancestry (B).

**Figure 3:**
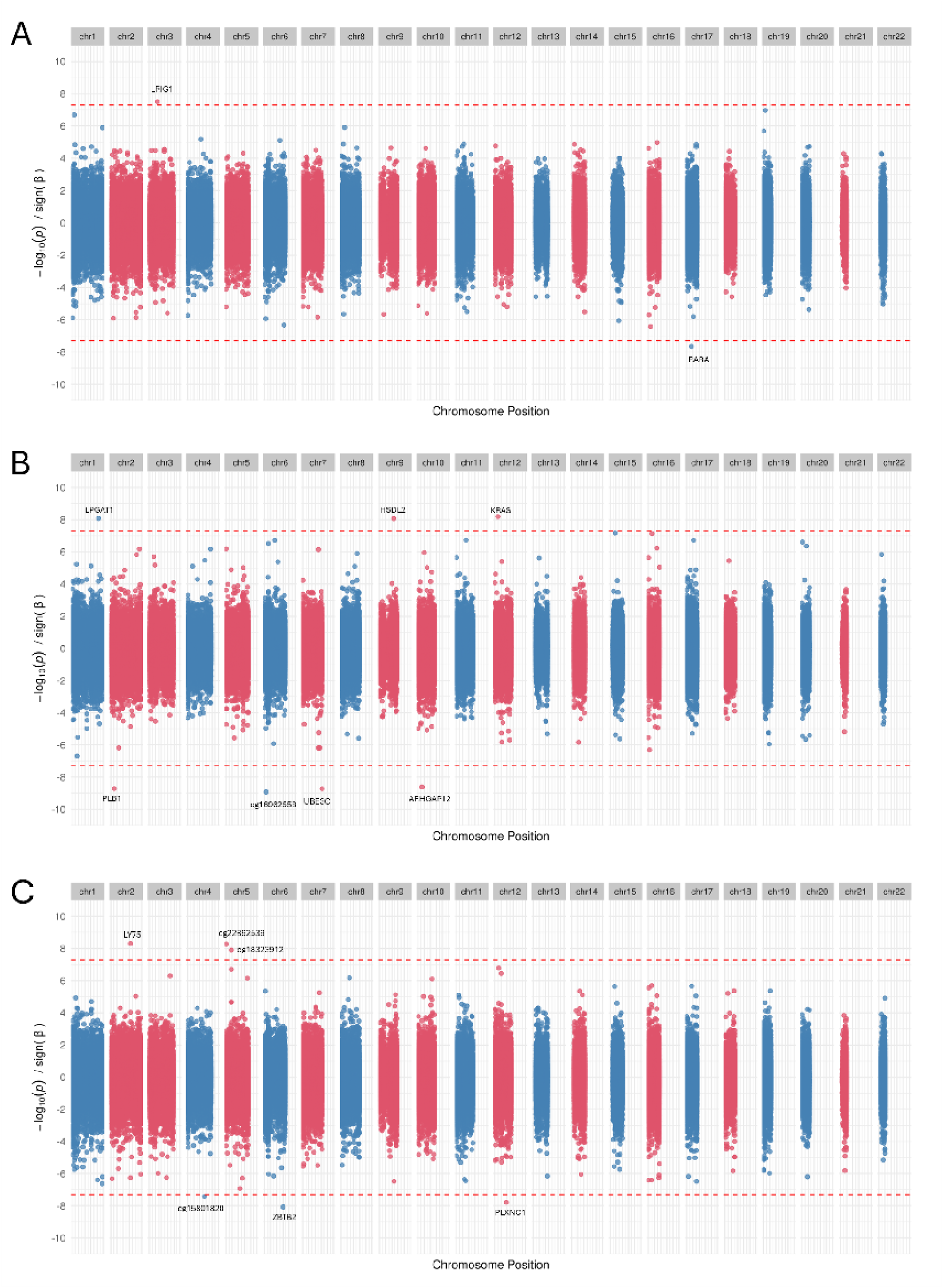
Sensitivity analysis. The Miami plots of sensitivity analysis EWAS performed using smoking and BMI information as additional covariates for (A) EUR, (B) AFR and (C) AMR ancestry.

In AMR, all 5 sites showing EWS in primary EWAS remained significant controlling for smoking and BMI. One additional site - cg15801820 on chromosome 4 showed a slightly lower P-value and became EWS in sensitivity analysis. Complete statistics of all significant CpG sites associations found in the sensitivity analysis for all 3 ancestries are provided in Table S1.

### EWAS in non-smokers

We then examined differential methylation at GAD-related CpG sites, focusing exclusively on non-smokers. Among the 26,099 individuals with available smoking data, 6,586 were identified as non-smokers, including 343 GAD cases and 6,243 controls. There were five CpG sites significantly associated with GAD (see Fig. S2). These sites correspond to the following genes: *DNAH10, SLC29A3, PTK2B, MAD2L2, and SSBP3*.

### Blood-brain DNAm correlation

We investigated DNAm changes in blood; a previous study by Braun *et al.* has shown a strong correlation between global DNA methylation levels in blood and brain CpG sites (rg=0.86) [22]. This suggests that blood DNAm levels may reflect brain DNAm levels. We evaluated blood-brain DNAm correlation of the 33 GAD associated sites in EUR using BECon [17]. Based on a study that included 16 samples using the Illumina 450□K Human Methylation Array, the BECon tool/database provides DNAm correlation information between blood and three different brain regions-Brodmann areas-BA7, BA10, and BA20 for a given CpG site. Of the 33 GAD-associated sites, BECon provided blood-brain correlation values for 14. Five of the 14 sites showed positive correlation while 9 sites showed negative correlation between blood methylation levels and at least one of the 3 BA brain regions. The absolute correlation values ranged between 0.1 to 0.6 (see Table S2). The highest correlations were observed for cg25648203 (Blood: BA10: −0.77) and cg19572487 (Blood: BA7 rho=0.64).

### Gene set enrichment analysis (GSEA)

GSEA was conducted to explore the potential enrichment of specific cell types or pathways in genes associated with CpG sites linked to GAD. The 33 CpG sites showing GAD association in EUR map to 18 unique genes. This gene set derived from all EUR EWAS revealed enrichment for the estrogen pathway, with a Bonferroni-corrected p-value (based on the number of gene sets included in the analysis) of 0.001 (see Fig S3). In the EUR sensitivity analysis, two genes with CpG sites passed the EWS threshold but did not show linkage to any common pathway/gene set. In the AFR EWAS, enrichment of *FOXP* related gene changes were observed (Fig S3). No pathway enrichment was observed in the gene set identified in AMR EWAS.

To capture more disease biology, we applied a more relaxed P-value threshold (P < 5e-05) to define the CpG site association and expand the gene set from the EUR EWAS results. With this relaxed cutoff, CpG sites in 91 and 367 genes showed associations with GAD in the sensitivity EWAS and all EUR EWAS respectively. No pathway enrichment was found in gene set from the sensitivity EWAS while 13 pathways including the previously identified estrogen signaling showed enrichment in the gene set derived from all EUR EWAS (Fig S3 & Supplementary Sheet Table 2). Enrichments of several chemical and genetic perturbation and chemical engineering based gene sets (from MSigDB) were also found. The gene set from sensitivity EWAS showed enrichment for 3 gene sets-related to RARA, FoxO and thyroid signaling (Supplementary Sheet). The relaxed cutoff-based gene set from all EUR EWAS also demonstrated enrichment for these three pathways, as well as additional pathways (Supplementary Sheet Table 3). There were no additional findings with AFR and AMR analysis.

**Table 3:** GAD nonEUR.

| Probe ID | CHR | Position | Coefficient | Coefficient SE | P-value | Gene |
| --- | --- | --- | --- | --- | --- | --- |
| **AFR** |  |  |  |  |  |  |
| cg11290603 | 1 | 2.12E+08 | 0.001137 | 0.000198 | 8.4315E-09 | LPGAT1 |
| cg09845761 | 2 | 28824172 | -0.00167 | 0.000297 | 2.1213E-09 | PLB1 |
| cg16962558 | 6 | 13512880 | -0.00383 | 0.000679 | 2.2372E-09 |  |
| cg25165171 | 7 | 1.57E+08 | -0.00269 | 0.000479 | 2.4636E-09 | UBE3C |
| cg20678641 | 9 | 1.15E+08 | 0.001336 | 0.00023 | 5.2884E- | HSDL2 |
|  |  |  |  |  | 09 |  |
| cg23663346 | 10 | 32198083 | -0.00149 | 0.000268 | 3.6139E-09 | ARHGAP12 |
| cg13532571 | 12 | 25404300 | 0.00142 | 0.000242 | 3.9781E-09 | KRAS |
| cg25271375 | 15 | 42565784 | 0.001542 | 0.000283 | 4.9738E-08 | GANC;TMEM87A |
| cg07038191 | 16 | 29801882 | 0.001322 | 0.000243 | 5.1984E-08 | KIF22 |
| **AMR** |  |  |  |  |  |  |
| cg24478096 | 2 | 1.61E+08 | 0.001551 | 0.000277 | 1.8071E-09 | LY75 |
| cg18323912 | 5 | 42994709 | 0.003508 | 0.000666 | 1.6323E-08 |  |
| cg22892539 | 5 | 1667258 | 0.013951 | 0.00254 | 3.7027E-09 |  |
| cg16222773 | 6 | 1.52E+08 | -0.00632 | 0.001251 | 1.1296E-08 | ZBTB2 |
| cg05136471 | 12 | 94577837 | -0.00348 | 0.000669 | 4.0361E-09 | PLXNC1 |

### Methylation risk scores

We investigated the performance of MRS in predicting GAD phenotype in EUR data. The total EUR cohort was split into discovery (75% samples) and validation cohorts (25% samples). With 75% of the EUR samples, the discovery EWAS found 6 EWS CpG sites for GAD (see table S4). Analogous to polygenic risk scores in GWAS studies, thresholding and pruning were used to select the CpG sites to be used in MRS calculation. Co-methylation signal was used to prune correlated sites for MRS. The MRS performed well at predicting GAD within MVP, showing a linear regression r-square value ranging from 0.018-0.02 (see Fig 4) with statistical significance for different P-value thresholds ranging from 0.005 to 5e-11. Complete details of MRS prediction regression results are shown in Table 4. Similar procedures were used to generate MRS in an independent cohort from Generation Scotland, using weights from the full MVP EWAS. Area under the receiver operating characteristic curve generated within the Generation Scotland ranged from 0.519 to 0.525, indicating that MRS performed at chance levels only and could not predict the GAD phenotype in this independent test sample.

**Figure 4:**
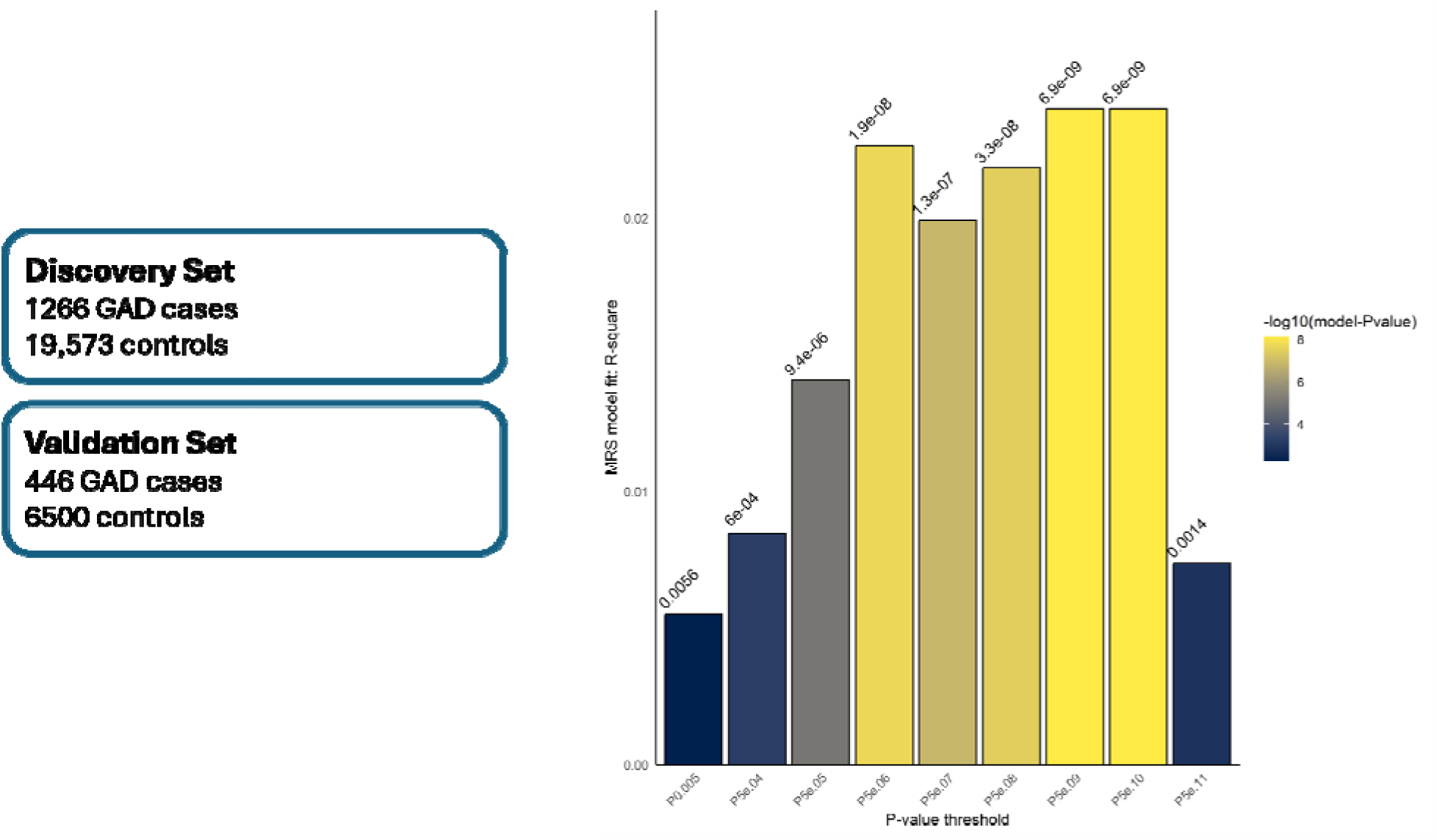
Methylation risk scores prediction in MVP. The left panel describes the discovery set and validation sets used to define CpG sites weights and predict GAD phenotype resp. The bar chart on right shows the performance of the linear regression models of MRS against the phenotype at different P-value thresholds used to consider the CpG sites to be used in MRS computation. The Y-axis shows the regression model r-square values at different P-value thresholds used shown on the x-axis. The color and P-value at the top of each bar depicts the significance -value of the regression model.

**Table 4:** Methylation Risk Scores Prediction.

| P-value thresh old | No. of CpG sites | No. Of independent CpG sites | *lm_R-square | lb95_R2 | ub95_R2 | lm_beta | lm_SE | lm_pval |
| --- | --- | --- | --- | --- | --- | --- | --- | --- |
| P5e.11 | 1 | 1 | 0.007371 | 0.001361 | 0.017429 | 0.009946 | 0.003105 | 0.001365633 |
| P5e.10 | 2 | 2 | 0.024019 | 0.008387 | 0.038638 | 0.018027 | 0.003108 | 6.89772619114135e-09 |
| P5e.09 | 2 | 2 | 0.024019 | 0.010726 | 0.045971 | 0.018027 | 0.003108 | 6.89772619114135e-09 |
| P5e.08 | 6 | 6 | 0.021843 | 0.007754 | 0.036849 | 0.017189 | 0.003108 | 3.33331299591484e-08 |
| P5e.07 | 14 | 14 | 0.019926 | 0.008915 | 0.038934 | 0.016402 | 0.003107 | 1.33769277450484e-07 |
| P5e.06 | 63 | 63 | 0.022654 | 0.010582 | 0.04094 | 0.017476 | 0.003103 | 1.85198316771304e-08 |
| P5e.05 | 240 | 238 | 0.01409 | 0.003776 | 0.030959 | 0.013763 | 0.003104 | 9.38608088011893e-06 |
| P5e.04 | 1054 | 1033 | 0.008459 | 0.000824 | 0.022367 | 0.010662 | 0.003106 | 0.000602573 |
| P0.005 | 5942 | 5764 | 0.005514 | 0.000438 | 0.017384 | 0.008601 | 0.003106 | 0.005632097 |

\*lm: linear regression model between PRS and GAD phenotype
lb95\_R2 and ub95\_R2: regression coefficients lower and upper bound at 96% confidence intervals

## Discussion

Epigenetic changes are a crucial bridge between genetic predispositions and environmental influences, potentially allowing us to uncover molecular alterations key genes mediating the biology of a phenotype. In this study, we conducted EWAS using methylation array data from 43,504 individuals (including 2,586 cases) from MVP cohort in three different genetic ancestry groups to decipher the epigenetic changes underlying GAD biology. In the largest ancestry – EUR EWAS, we identified differential methylation status of 33 CpG sites across 18 genes.

Implicated genes include *RARA, LRIG1, FURIN, KRAS, GCNT2,* and *F2RL3*. *RARA* (retinoic acid receptor alpha) encodes a nuclear receptor and key regulatory component in retinoic acid (RA) signaling. Previous knockdown studies in mice have demonstrated the role of RA signaling in the brain in modulating emotional behavior, influencing anxiety-like behavior and depression-like traits [23]. We identified EWS association between methylation of CpG sites cg19572487 and cg17739917 in the gene encoding *RARA* and GAD, further supporting these findings, suggesting that RARA may be a crucial regulatory gene in mediating the effects of RA signaling manipulations in GAD. The LRIG1 (leucine rich repeat and immunoglobulin like domain protein 1) is a tumor suppressor gene expressed in stem cells in brain, colon and skin [24]. It inhibits receptor tyrosine kinase (RTK) signaling and acts as a gatekeeper to exit quiescence in adult neural stem cells. In aging mouse brain hippocampus, LRIG1 overexpression has been shown to block neurotrophin signaling and negatively impact the development and integration of adult borne granule cells, resulting in abnormal dendritic development and impaired cognitive function [25]. The strong association of CpG site - cg16414530 methylation in *LRIG1* gene with anxiety suggests that the *LRIG1*-mediated abnormal neurogenesis and development may contribute to the pathophysiology of anxiety. This CpG site may serve as a salient marker for understanding epigenetic mechanisms underlying this important regulatory protein. *FURIN* is another important gene known to be associated with various psychiatric disorders like schizophrenia, cannabis and opioid use disorders, bipolar affective disorder, and major depressive disorder. Studies have reported reduced levels of *FURIN*’s substrate protein - BDNF in brain in anxiety-like behaviors [26], suggesting the potential involvement of *FURIN* in GAD pathophysiology. The association of methylation levels at multiple CpG sites (see Table 2) in *FURIN* provides evidence to this link at the epigenetic level. Additionally, *GCNT2* and *SUGT1P1* exhibit CPG sites associated with GAD. A previous EWAS study. identified hypomethylation at CpG site cg05157878 in *GCNT2* and at cg13920529 site in *SUGT1P1* to be associated with childhood abuse and neglect, including emotional trauma [33]. Overall, our findings indicate that many of the 18 identified genes have established links to psychiatric disorders, with some specifically linked to anxiety and anxiety disorders. Furthermore, the identification of 33 sites highlights key regulatory regions within these genes, providing potential targets for further research.

Factors, such as smoking or BMI, are known to influence DNA methylation patterns [20, 21]. In an EWAS study, this creates the potential for confounding, where methylation signals related to the phenotype of interest may overlap with those from these additional environmental factors. To separate the methylation signal specifically associated with GAD from the influence of these environmental factors, we conducted a sensitivity analysis, using smoking and BMI data to regress out their effects on GAD. The sensitivity analysis confirmed two CpG sites—cg17739917 on the *RARA* gene and cg16414530 on the *LRIG1* gene—as specific to GAD. The remaining 31 sites do not survive the epigenome wide significance threshold (P. value < 5.88e-08) following this correction for smoking and BMI. This may be due to the smaller effect size or stringent threshold, limited sample size, the complexity of gene-environment interactions, or the reduction in power that this analysis entailed. Many of these sites are located in genes like FURIN, F2RL3 which are linked to psychiatric and neurodegenerative disorders, further suggesting their potential relevance to GAD pathology despite not surviving correction for smoking status [26, 34]. Further research is needed to confirm whether these sites are GAD-specific or influenced by environmental factors or a result of both.

Fourteen of the 33 identified CpG sites have previously shown correlated methylation patterns in both blood and brain, suggesting that the GAD linked sites identified in this work are likely to have similar GAD-associations in brain as well. These 14 sites, therefore, may represent high-priority candidate CpG sites for future experimental and animal model studies aimed at understanding the brain signaling alterations underlying GAD biology.

In the EUR EWAS analysis, the 18 EWS GAD-linked genes with CpG site associations showed enrichment in the estrogen signaling pathway gene set. (Fig S3 & Supplementary Sheet1). The genes from AFR EWAS revealed a similar enrichment of FOXP related gene changes (Fig S3). FOXP1 is an estrogen inducible transcription factor and estrogen pathway. Prior work has revealed evidence for hypermethylation across five CpG sites in promoters and enhancers of FOXP3 in female patients with diagnosed panic disorders [35]. These findings are consistent with previous research, which has also implicated estrogen and FOXP signaling and in the biology of GAD, including in studies of risk variants [11, 36–39]. We previously found genetic variations near the gene for estrogen receptor 1 (ESR1) to be related to GAD symptoms in a large GWAS [11].

To maximize discovery of biological signal related to GAD (at the risk of increasing false positives), we applied a relaxed P-value threshold (P-value < 5e-05) to the EUR EWAS and used the resulting finding to perform GSEA(see Supplementary Sheet). The relaxed cutoff revealed the enrichment of GAD-linked genes for 3 more gene sets related to FoxO, RARA, thyroid signaling. These enrichments were consistent in both all EUR and in the follow-up sensitivity analyses for smoking and BMI. Abnormal thyroid signaling and anxiety are often associated due to overlapping symptoms (nervousness, restlessness, palpitations, irritability), influence on common neurotransmitters (GABA receptors) and altered T4 levels in anxiety and heightened stress response in both [40–43]. There have been reports of involvement of FoxO signaling in the stress response and anxiety as well [44]. Further research on the epigenetic alterations of these pathways in GAD can enhance our understanding of how disruptions in these pathways influence the development and progression of GAD.

This study has some limitations. Our study was cross-sectional, with a mean age of blood draw at approximately 68. As the age of onset for GAD is approximately between 21 and 35 [45], some time would have passed for many case participants between initial diagnosis and measurement of methylation levels by array. Conversely, controls will have traversed the typical risk age range for diagnosis and are less likely to be false negatives. It remains to be determined how much age at measurement may influence our results. Additionally, while our study provides a strong foundation for understanding the GAD methylome signaling through a large sample size EWAS, the current data derived from GAD-linked methylation signal-driven gene sets remains limited. Testing this MRS into an independent cohort from Generation Scotland using the AUC of the receiver operating characteristic revealed performance similar to chance. To identify more meaningful pathway alterations related to GAD, additional research with large sample sizes revealing more GAD-linked genes is crucial. Additionally, this sample draws from a population of US Veterans receiving care from the Veterans Affairs hospital system, and thus is mostly male.

In conclusion, we conducted a large-scale epigenetic analysis of GAD using data from the MVP, stratified by three genetic ancestries: EUR, AFR, and AMR. Our findings identified 49 CpG sites reaching epigenome-wide significance for GAD across these populations. However, when accounting for smoking—either by covarying for smoker status or restricting analyses to non-smokers—the number of significant CpG sites was reduced from 33 in the full EUR analysis to 2, highlighting the potential confounding effect of smoking on epigenetic associations. Although we had fewer samples of non-EUR ancestry, these populations did not exhibit as much attenuation in signal when covarying by smoking status. Additionally, we introduced a novel approach for constructing methylation risk scores, drawing from threshold and clumping methodologies in genetic studies. This method aims to mitigate bias by leveraging co-methylation patterns to refine risk score calculation. We assessed the performance of methylation risk scores at predicting GAD phenotype in the European cohort and found that the MRS captured ∼2% of the variance in the GAD phenotype. Our findings contribute to a deeper understanding of the epigenetic architecture of GAD and provide a framework for future studies aiming to integrate epigenetic risk profiling into psychiatric genetics.

## Supporting information

MVP Core Acknowledgement

## Data Availability

Data access will be provided through VA Cipher following acceptance of the manuscript.

## Acknowledgement

This research is based on data from the Million Veteran Program, Office of Research and Development, Veterans Health Administration, and was supported by MVP000, MVP092 and MVP069 as well as awards #2IO1BX006482, 5R01MH133728-02, and #5IK2BX005058. This publication does not represent the views of the Department of Veteran Affairs or the United States Government.

