## Supplement for "Multi-ancestry epigenome wide association study of generalized anxiety disorder"

**Supplementary Data**

**Supplementary Tables**

Table S1: Significant GAD-associations in the sensitivity analysis of EWAS for all three ancestries.

|  | **Probe ID** | **CHR** | **Position** | **Coefficient** | **Coefficient SE** | **P value** | **Gene** |
| --- | --- | --- | --- | --- | --- | --- | --- |
| **EUR** |  |  |  |  |  |  |  |
|  | cg16414530 | chr3 | 66437745 | 0.00445 | 0.000846 | 3.09764737104338e-08 | LRIG1 |
|  | cg17739917 | chr17 | 38477572 | -0.00703 | 0.001425 | 2.1913300145221e-08 | RARA |
| **AFR** |  |  |  |  |  |  |  |
|  | cg25165171 | chr7 | 1.57E+08 | -0.00269 | 0.000488 | 1.854902314156e-09 | UBE3C |
|  | cg23663346 | chr10 | 32198083 | -0.00149 | 0.000272 | 2.42088165629699e-09 | ARHGAP12 |
|  | cg20678641 | chr9 | 1.15E+08 | 0.00133 | 0.000234 | 8.48017422724254e-09 | HSDL2 |
|  | cg16962558 | chr6 | 13512880 | -0.00383 | 0.000686 | 1.17650369901914e-09 |  |
|  | cg13532571 | chr12 | 25404300 | 0.001415 | 0.000247 | 6.42094494217618e-09 | KRAS |
|  | cg11290603 | chr1 | 2.12E+08 | 0.001145 | 0.000202 | 8.23573792897602e-09 | LPGAT1 |
|  | cg09845761 | chr2 | 28824172 | -0.00167 | 0.000302 | 1.88743650266425e-09 | PLB1 |
| **HIS** |  |  |  |  |  |  |  |
|  | cg24478096 | chr2 | 1.61E+08 | 0.001536 | 0.000284 | 4.96738673580664e-09 | LY75 |
|  | cg22892539 | chr5 | 1667258 | 0.013929 | 0.002583 | 5.38712424519382e-09 |  |
|  | cg18323912 | chr5 | 42994709 | 0.003555 | 0.000675 | 1.24404034354903e-08 |  |
|  | cg16222773 | chr6 | 1.52E+08 | -0.00641 | 0.001266 | 8.36454836610044e-09 | ZBTB2 |
|  | cg15801820 | chr4 | 1.38E+08 | -0.00748 | 0.00155 | 3.73725969704535e-08 |  |
|  | cg05136471 | chr12 | 94577837 | -0.00344 | 0.000695 | 1.62233112673845e-08 | PLXNC1 |

Table S2: Blood-brain correlations of methylation levels at GAD associated sites found in EUR EWAS

| **CpG ID** | **Chr (hg19)** | **Associated Genes** | **CpG in Feature of Gene, respectively** | **Cor Blood-BA7** | **Cor Blood- BA10** | **Cor Blood- BA20** |
| --- | --- | --- | --- | --- | --- | --- |
| cg05575921 | 5 | AHRR | intragenic | -0.026 | **-0.394** | -0.321 |
| cg25648203 | 5 | AHRR | intragenic | -0.406 | **-0.776** | -0.35 |
| cg02716826 | 9 | AQP3 | intragenic | -0.059 | **-0.171** | 0.144 |
| cg00501876 | 3 | CSRNP1 | intragenic | **-0.368** | -0.188 | -0.006 |
| cg03636183 | 19 | F2RL3 | intragenic | **-0.406** | -0.018 | -0.074 |
| cg05284742 | 14 | ITPK1 | intragenic | 0.103 | **-0.253** | 0.109 |
| cg16552271 | 1 | KAZN | intragenic | **0.397** | 0.247 | -0.359 |
| cg09182189 | 1 | NADK | intragenic | 0.232 | -0.176 | **-0.465** |
| cg04885881 | 1 | intergenic | intergenic | 0.182 | **0.303** | 0.009 |
| cg24859433 | 6 | intergenic | intergenic | 0.029 | **0.112** | 0.056 |
| cg15342087 | 6 | intergenic | intergenic | **0.215** | 0.153 | 0.138 |
| cg01940273 | 2 | intergenic | intergenic | **0.368** | 0.226 | 0.088 |
| cg11660018 | 11 | PRSS23 | promoter | **-0.509** | -0.168 | -0.341 |
| cg19572487 | 17 | RARA | intragenic | **-0.641** | -0.271 | -0.059 |

**Supplementary Figures**

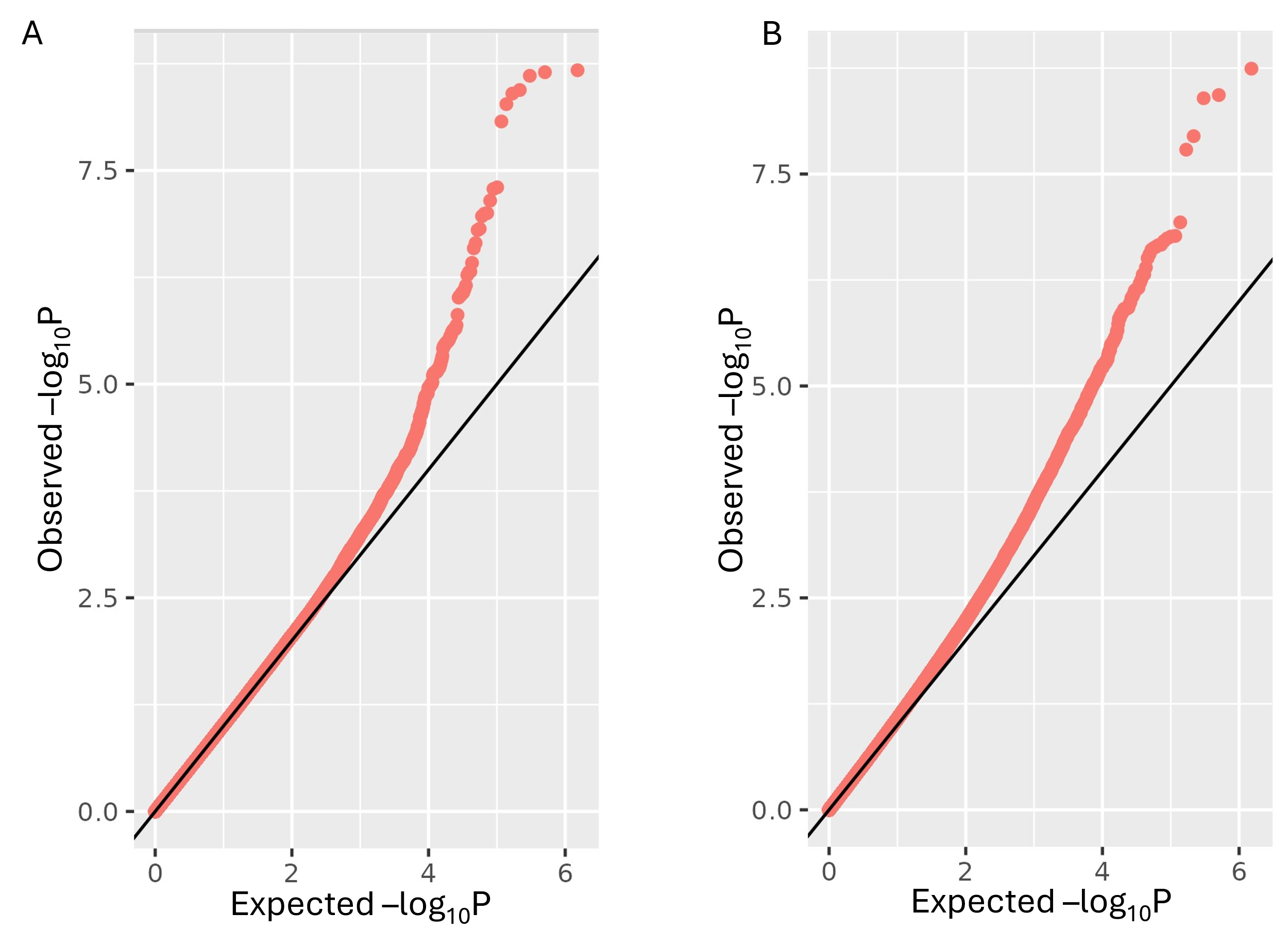

Figure S1: QQ plots of GAD EWAS in (A) AFR & (B) Admixed cohort. The inflation factor value in AFR EWAS is 1.02 and in Admixed EWAS is 1.09.

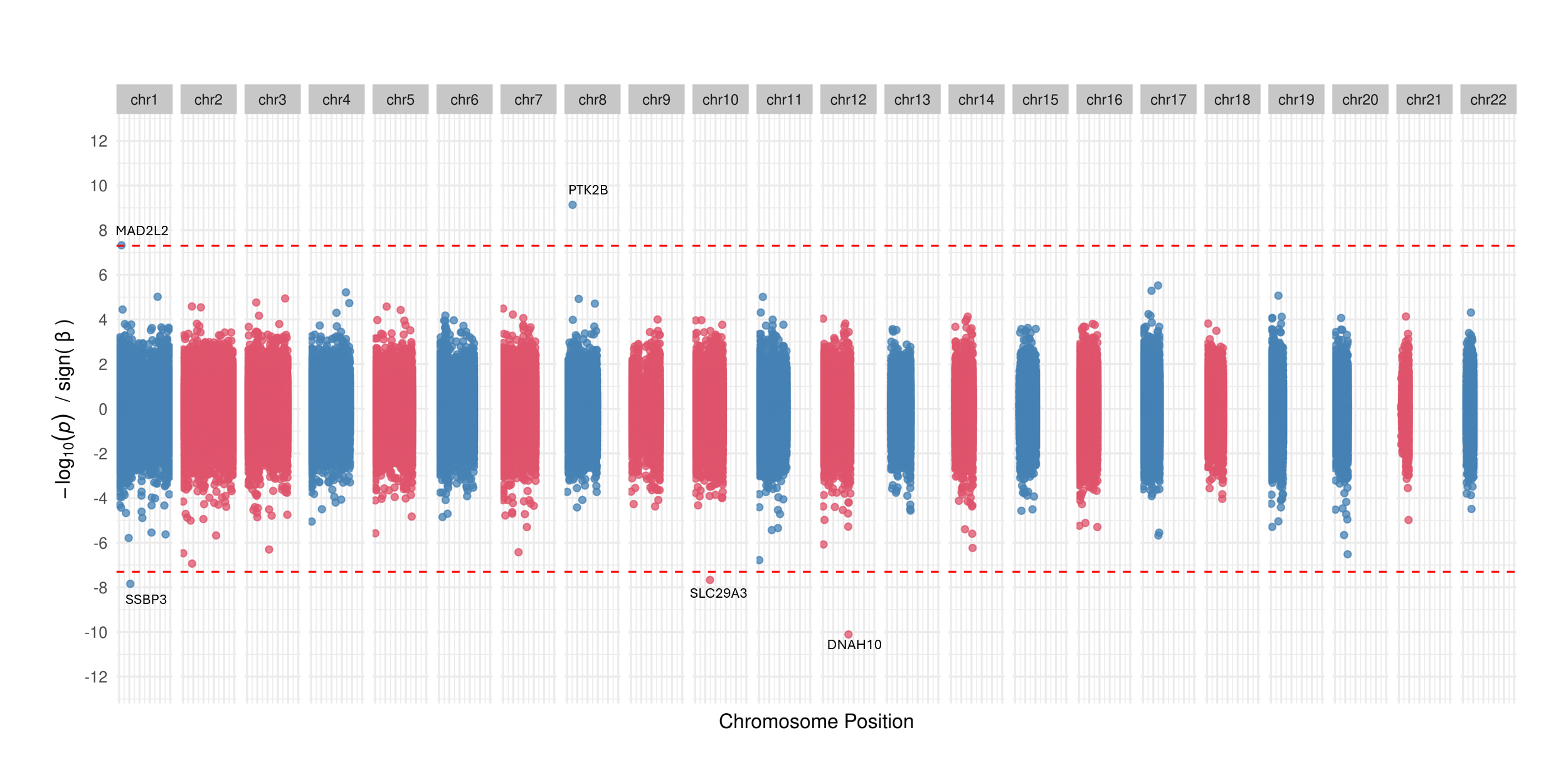

Figure S2: Miami plot showing the GAD associated CpG sites found in EWAS performed in non-smokers sample.

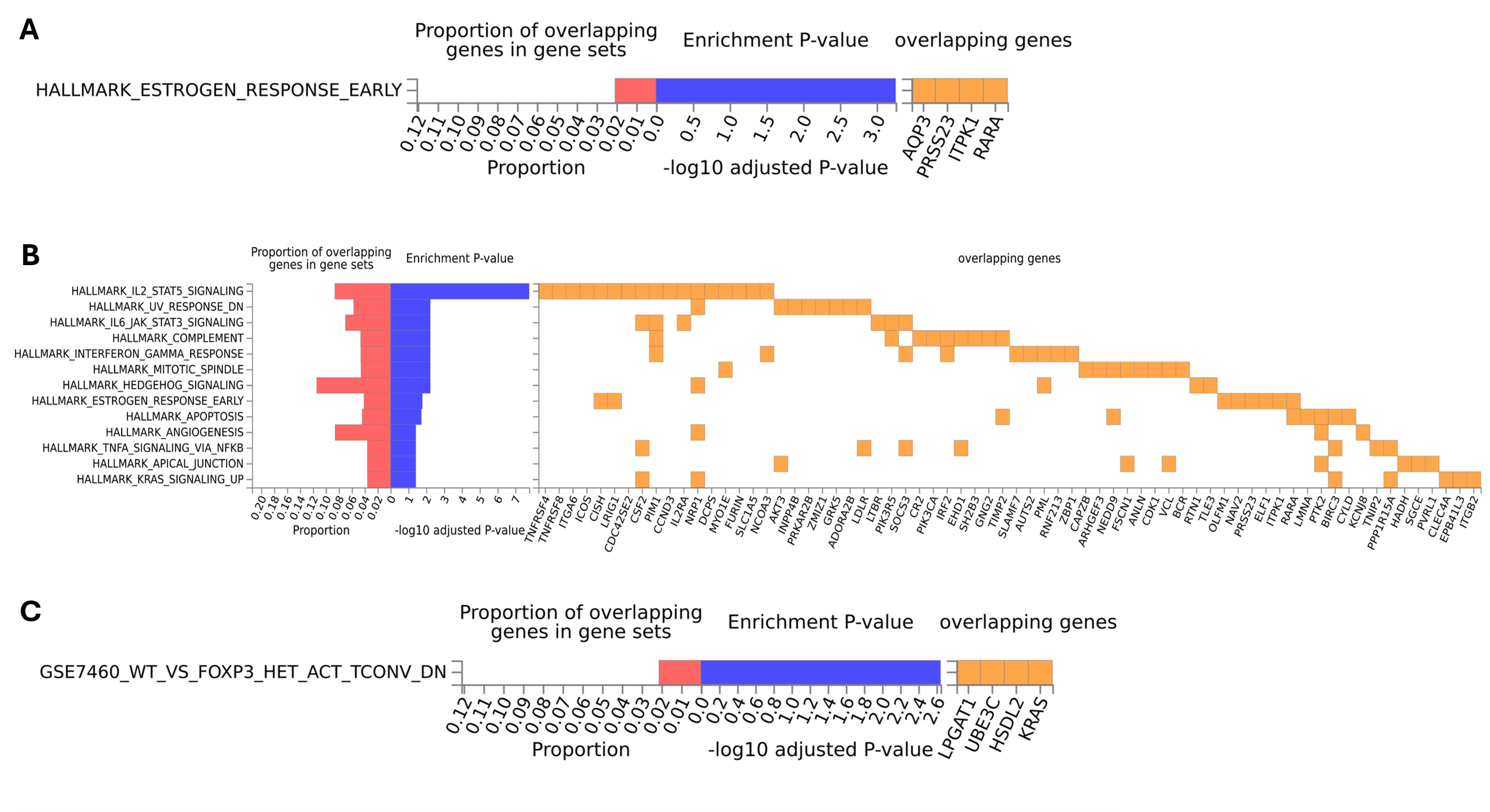

Figure S3: Gene set enrichment results. A, B shows the statistics of pathway (MSigDB hallmark pathways) found to be enriched in the genes derived from EUR EWAS with EWS threshold (A) & with relaxed threshold (B). (C) shows the statistics of pathway enriched in the genes mapping to 9 EWS CpG sites from AFR EWAS

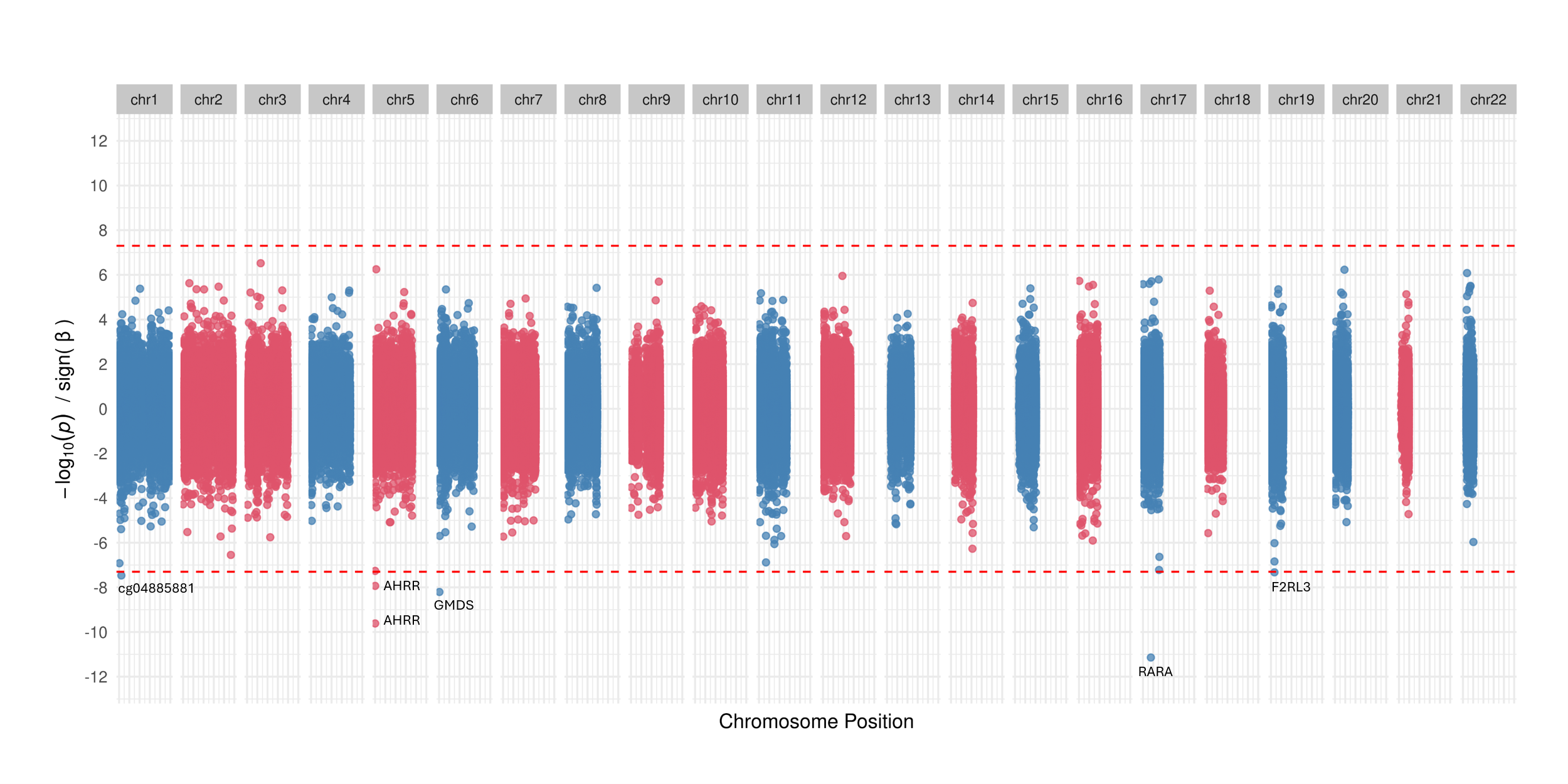

Figure S4: Miami plot showing the CpG site associations found in the EWAS performed in discovery cohort.
